# Coating formulation change is the root cause for inferior bioefficacy of long-lasting insecticidal nets in Papua New Guinea

**DOI:** 10.1101/2022.06.25.22276903

**Authors:** Nakei Bubun, Evodia Anetul, Melanie Koinari, Tim Freeman, Stephan Karl

## Abstract

In a study published in Nature Communications in August 2020, we demonstrated an abrupt decrease in the bioefficacy of long-lasting insecticidal nets (LLINs) for malaria prevention delivered to Papua New Guinea (PNG) between 2012 and 2013. This coincided with a rise in malaria cases in the country. At the time of publication of the original article, we were unable to pinpoint the exact reasons for the observed shift towards inferior product performance and stated that ‘further studies are required to determine the underlying cause of the observed reduced bioefficacy of these LLINs’ ^1^. Due to the potentially significant public health implications (hundreds of millions of this specific LLIN product had been distributed globally), our study led to discussions and speculation among stakeholders. Here, we present data unequivocally showing that the observed reduction in the ability to kill mosquitoes of these LLINs is a direct result of a manufacturing change that occurred at the same time.

## Main Text

Long-lasting Insecticidal Nets (LLINs) are the most important vector control tool against malaria ^2^. No other method is considered to have prevented more cases and saved more lives ^3^. LLINs protect by providing a physical barrier between the user and potentially infectious mosquitoes. Equally importantly, they also afford community protection through efficiently killing mosquitoes that come into contact with the insecticide-treated surfaces ^4^. The vast majority of LLINs is distributed with public donor funding and undergoes a WHO prequalification process ^5^.

In our original study, we described that post-delivery testing of LLINs in Papua New Guinea (PNG) revealed a significant and abrupt drop in the ability of LLINs to kill susceptible colony mosquitoes between 2012 and 2013. This was surprising as the same LLIN product had been distributed in PNG since the beginning of the mass distribution campaigns in 2007 until 2019 and thus, consistent product performance was assumed.

PNG is the country with the highest malaria transmission outside of Africa. The distribution of the significantly less potent nets in PNG coincided with a resurgence of malaria in the country and we hypothesised that the decreased community-level protection afforded by these inferior LLINs had contributed to the observed increase in malaria.

However, it has remained subject to speculation what had caused the performance of these LLINs to be so dramatically reduced. Hypotheses to explain our observations included inappropriate transport, and short-term storage conditions of the new and unused nets after 2013, such as exposure to elevated temperature in shipping containers ^6^. We showed that this was unlikely, as the overall insecticide content in the tested nets from all years (2007 to 2019) was very similar, i.e., container storage had not resulted in a rapid breakdown of the insecticide ^7^. Also, older nets that had been stored for much longer and exhibited 100% kill rate had, on average, slightly lower insecticide content which we attributed to natural decay over many years of storage ^7^. In addition, we showed that short-term heating of the LLINs in question increased their potency to kill mosquitoes (rather than to decrease it), which we attributed to a heat-facilitated migration of the insecticide from inside the coating to the net surface ^8,9^.

It was also suspected that the mosquito strain that we had used (a pyrethroid susceptible strain of *Anopheles farauti*) or technicalities related to conducting WHO cone bioassays at the PNG Institute of Medical Research were responsible for the observed inferior performance. We ruled out these possibilities categorically, by conducting a multi-centre trial with the same nets that showed that our observations from PNG were reproducible in an African ‘Good Laboratory Practice’ accredited facility ^10^.

A third hypothesis was that the observed abrupt shift from very potent killing of mosquitoes to inferior insecticidal performance between 2012 and 2013 was due to a manufacturing change that had occurred at the same time and that was not detectable in routine LLIN predelivery inspections. Initial analyses using X-ray fluorescence spectrometry had hinted towards this possibility ^5^ but a definitive conclusion remained elusive.

The LLIN product that was distributed in PNG between 2007 and 2019 (PermaNet® 2.0) is a polyester net with a polymer coating that contains the insecticide. While predelivery inspections and our own work had verified the total insecticide content of all nets to be within specifications ^5^, the formulation of the polymer coating may also influence insecticidal potency of a net ^9^. This is because some coating technologies and formulations more effectively present the insecticide on the net surface where it comes into contact with mosquitoes, whereas other coatings can enclose the insecticide under a polymer layer, essentially ‘locking it in’ and restricting its bioavailability.

Coatings in the textile industry can be grouped into a few major classes, with a major distinction between polyfluorocarbon-based (PFC) and non-PFC coatings. Non-PFC coatings are typically acrylates, polyurethanes or mixtures thereof. These are cheaper and considered more environmentally friendly.

The industrial standard to distinguish between PFC and non-PFC formulations is the detection of fluorine in the coating polymer as it is only found in PFC coatings. The reference method to measure total polymer fluorine content is combustion ion chromatography, where a sample is incinerated in an oxygen bomb and the released fluorine is measured using ion chromatography.

To test the hypothesis of a major coating technology change, we submitted a total of n=12 nets for combustion ion chromatography to be conducted by an independent, globally recognised reference laboratory (SGS, Sydney, Australia). Specifically, the tested samples were from LLINs also used in the original study from the following years: 2008 (n=2), 2010 (n=2), 2012 (n=2), 2015 (n=2), 2017 (n=2) and 2019 (n=2). Given that the change in the bioefficacy of the nets occurred after 2012, we expected the first 6 nets (2008-2012) to be coated using one specific coating technology, whereas the other 6 nets (2015-2019) would be coated with another coating technology.

The independent analysis unequivocally proved this hypothesis. The nets from before and including the year 2012 (n=6) contained high amounts of fluorine on the order of 3-4 g/kg, which corresponds well to the expected weight ratio of the coating itself. The nets after 2012 (n=6) contained only small trace amounts of fluorine (approx. 0.02 g/kg) as shown in Figure 1.

**Figure 1:**
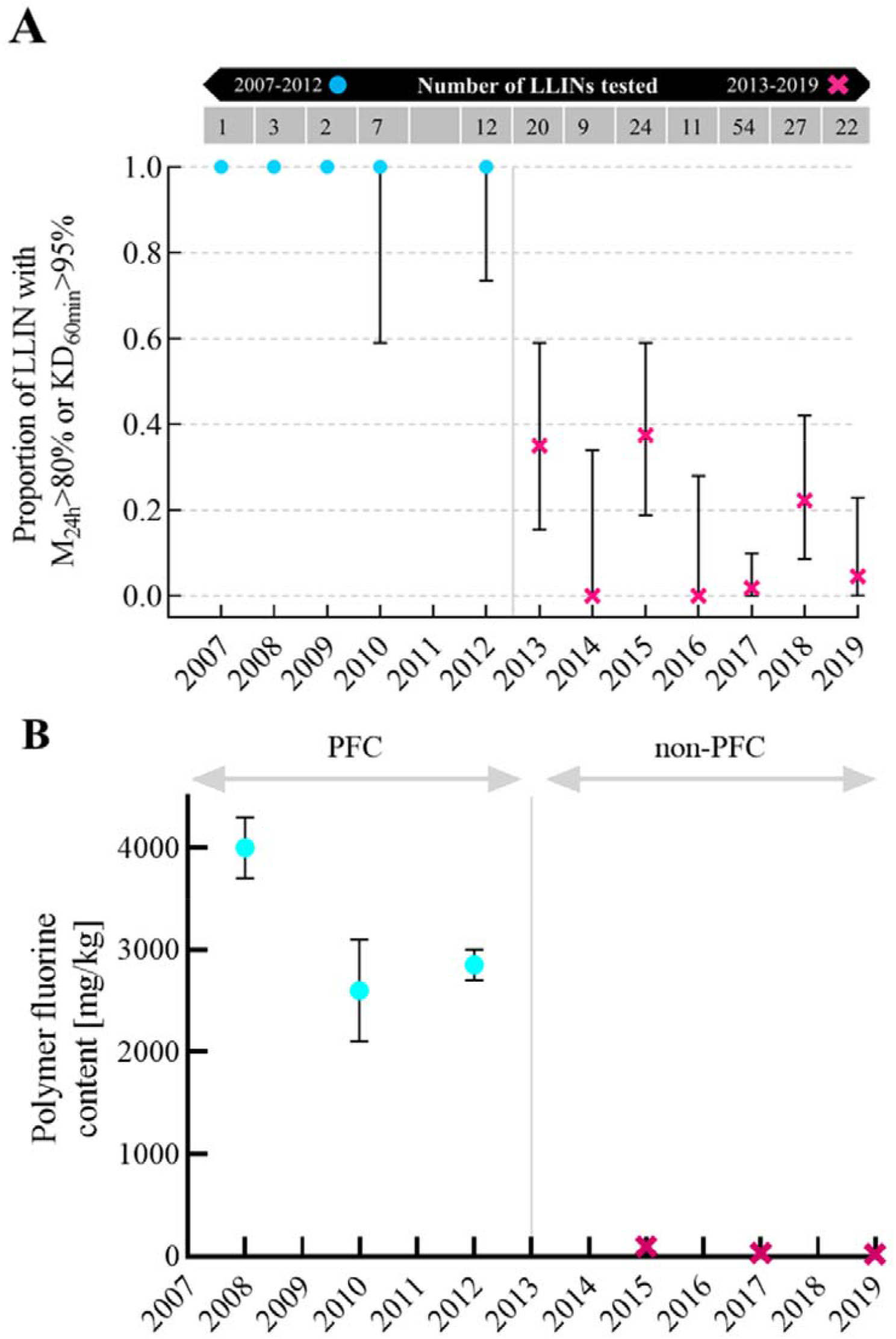
Change in coating technology from PFC to non-PFC resulted in decreased bioefficacy. A) Decreased bioefficacy of PermaNet® 2.0 as presented in the original study [2]. B) Corresponding polymer fluorine content as determined by combustion ion chromatography in the present study on samples from the same nets.

Thus, it is without question that the coating technology in the nets that killed mosquitoes very effectively (pre-2012) and those that lack this ability (post-2012) is very different. While nets pre-2012 where coated with PFC technology, the nets manufactured after 2012 were coated with a non-PFC formulation, leading to a completely altered LLIN product with a significantly inferior ability to kill susceptible *An farauti* mosquitoes.

One may hypothesise that if the insecticide is enclosed under a layer of polymer coating, a single wash may remove that layer and make the inferior nets perform normally. To test this, we conducted standard WHO wash assays (Figure 2). Our analyses show that post-2012 PermaNet® 2.0 LLINs distributed in PNG performed poorly over their entire lifespan (up to 25 washes) as shown in Figure 2.

**Figure 2:**
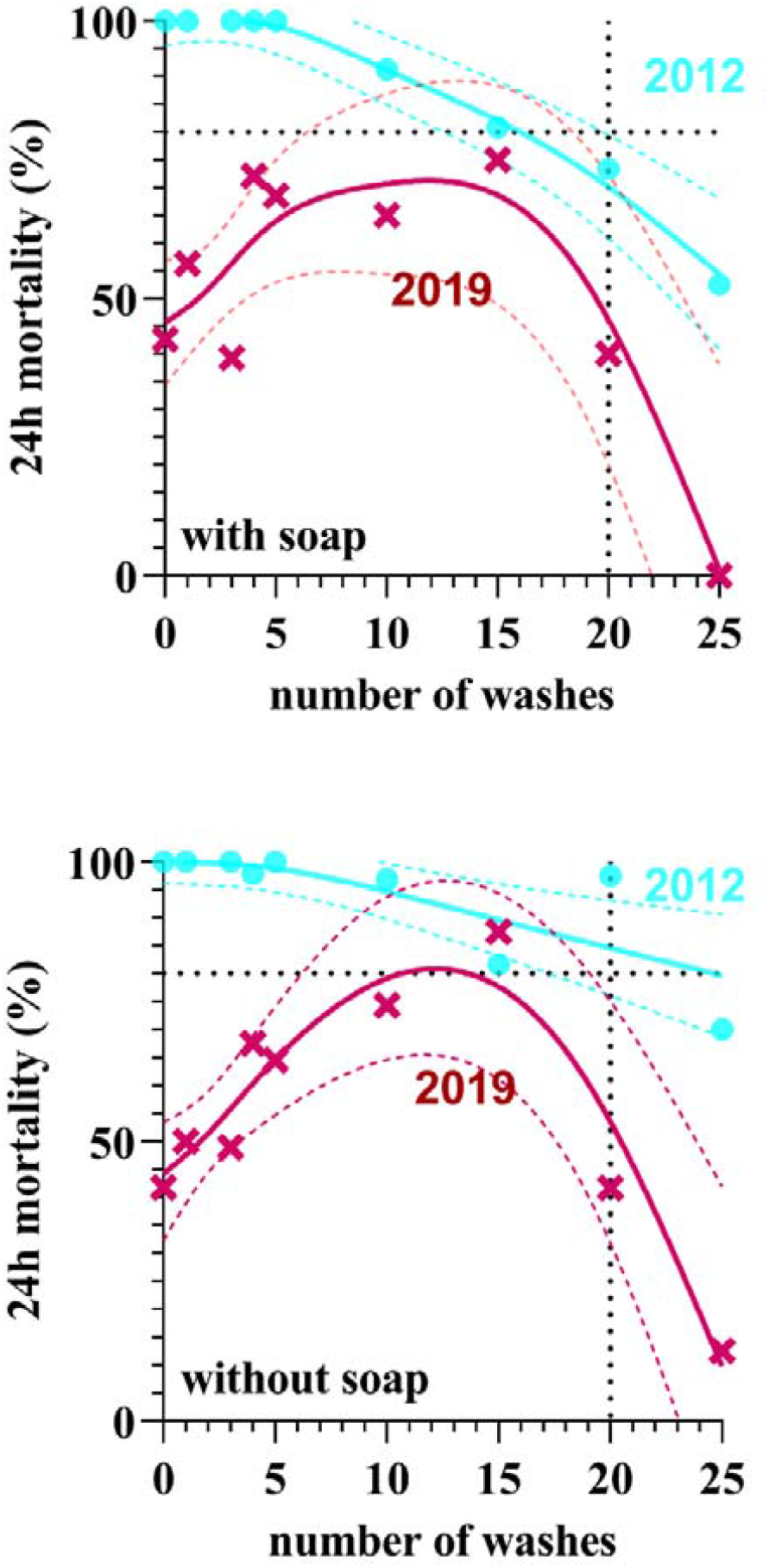
Standardised WHO Cone Bioassay results after washing. A) shows cone bioefficacy data (24h mortality) for PermaNet® 2.0 nets LLINs from 2012 (n = 2 nets, 7 peices per net) and LLINs from 2019 (n = 2, 7 pieces per net) washed according to WHO guidelines using a local soap. B) To better understand the effect of the soap, we also conducted the same wash tests on the nets with water only, with no soap added at all.

In conclusion, our work demonstrates that coating technology of LLINs is an essential product attribute that determines how effectively these essential public health commodities kill malaria mosquitoes. LLIN coatings thus need to be controlled and any change to them needs to be validated to not detrimentally affect insecticidal performance. PermaNet® 2.0 was the most widely distributed LLIN product in the world at the time. Given that LLIN coating formulations have not been controlled or regulated since mass distributions began, it is likely that many LLIN manufacturers have changed coating formulations over the years. While it is not possible to exactly quantify the number of malaria cases that could have been averted if this manufacturing change had not been made, we consider it highly likely that it has contributed to the stalling success in malaria case reduction across the world, and especially in countries that have relied solely on PermaNet® 2.0, like PNG.

## Data Availability

All data produced in the present study are available upon reasonable request to the authors.

https://bit.ly/3yV07F7

## Author contributions statement

Conceived study: SK, TWF; Conducted Experiments and processed samples: NB;EA;MK

## Supporting Information File

Total Fluorine Polymer Content: Official Report from SGS

