## Supporting Information File for "Coating formulation change is the root cause for inferior bioefficacy of long-lasting insecticidal nets in Papua New Guinea"

### ANALYTICAL REPORT

| | |
|---|---|
| Client: | Stephan Karl<br>**James Cook University**<br>88 Macgregor Rd<br>Smithfield QLD 4878 |
| Your Reference: | Long Lasting Insecticidal Net for Total Fluorine Analysis |
| SGS Report Number: | SP035171 |
| Date of Receipt of Samples: | 09/05/2022 |
| Sample Description: | 12 net samples individually wrapped in aluminium foil |

The samples were analysed in accordance with your instructions. The results and associated information are contained in the following pages of the report. Should you have any queries regarding this report please contact the undersigned.

Reported by: Dr Tracey Yeung

Date: 18/05/2022

Report authorised by: Dr Peter Novella

Date: 18/05/2022

This document is issued, on the Client's behalf, by the company under its General Conditions of Service available on request and accessible at http://www.sgs.com/terms_and_conditions.htm. The client's attention is drawn to the limitation of liability, indemnification and jurisdiction issues defined therein.

Any other holder of this document is advised that information contained hereon reflects the company's findings at the time of its intervention only and within the limits of client's instructions, if any. The company's sole responsibility is to its client and this document does not exonerate parties to a transaction from exercising all their rights and obligations under the transaction documents.

This test report shall not be reproduced except in full, without written approval of the laboratory.

#### Background:

SGS Sydney was requested by Stephan Karl of James Cook University to analyse 12 net samples for total fluorine. The samples were received on 09/05/2022 and logged in with the following laboratory reference numbers:

**Table 1: Sample Information**

| SGS ID | Client ID | Year of Manufacture |
|---|---|---|
| SP035171-1 | PermaNet 2.0<br>Long Lasting Insecticidal Net 1 | 2008 |
| SP035171-2 | PermaNet 2.0<br>Long Lasting Insecticidal Net 2 | 2008 |
| SP035171-3 | PermaNet 2.0<br>Long Lasting Insecticidal Net 3 | 2010 |
| SP035171-4 | PermaNet 2.0<br>Long Lasting Insecticidal Net 4 | 2010 |
| SP035171-5 | PermaNet 2.0<br>Long Lasting Insecticidal Net 5 | 2012 |
| SP035171-6 | PermaNet 2.0<br>Long Lasting Insecticidal Net 6 | 2012 |
| SP035171-7 | PermaNet 2.0<br>Long Lasting Insecticidal Net 7 | 2015 |
| SP035171-8 | PermaNet 2.0<br>Long Lasting Insecticidal Net 8 | 2015 |
| SP035171-9 | PermaNet 2.0<br>Long Lasting Insecticidal Net 9 | 2017 |
| SP035171-10 | PermaNet 2.0<br>Long Lasting Insecticidal Net 10 | 2017 |
| SP035171-11 | PermaNet 2.0<br>Long Lasting Insecticidal Net 11 | 2019 |
| SP035171-12 | PermaNet 2.0<br>Long Lasting Insecticidal Net 12 | 2019 |

#### Methods Used:

A portion of each sample was completely combusted in an oxygen bomb. The resulting combustion products were diluted with water (Type I) and the concentration of fluorine was analysed by ion chromatography.

#### Analytical Results:

**Table 2: Total Fluorine Analysis Results**

| SGS ID | Fluorine | Units |
|---|---|---|
| SP035171-1 | 4300 | mg/kg |
| SP035171-2 | 3700 | mg/kg |
| SP035171-3 | 2100 | mg/kg |
| SP035171-4 | 3100 | mg/kg |
| SP035171-5 | 3000 | mg/kg |
| SP035171-6 | 2700 | mg/kg |
| SP035171-7 | 120 | mg/kg |
| SP035171-8 | 68 | mg/kg |
| SP035171-9 | 37 | mg/kg |
| SP035171-10 | 24 | mg/kg |
| SP035171-11 | 35 | mg/kg |
| SP035171-12 | 5.2 | mg/kg |

#### Opinions and Interpretations: N/A

SGS Australia Pty Ltd
ABN 44 000 964 278
Environment, Health & Safety Unit 16, 33 Maddox St Alexandria 2015 NSW Australia
t +61 (0)2 8594 0400 f +61 (0)2 8594 0499 www.au.sgs.com
